# Automated Risk Assessment of Amputation in Patients with Peripheral Artery Disease

**DOI:** 10.64898/2025.12.02.25340994

**Authors:** Marko Milosevic, Tanmay Demble, Christine Lary, Elsie Gyang Ross, Saeed Amal

## Abstract

**Objective:** Peripheral artery disease (PAD) affects over eight million Americans and is a leading cause of non-traumatic lower extremity amputation in the United States. Identifying which patients are at highest risk for limb loss can improve the timeliness of intervention to prevent amputation. We wanted to test the predictive performance of machine learning models developed to predict amputations in patients with peripheral artery disease (PAD) at various intervals of time (60, 120, 180, and 365 days prior to amputation) and across two different clinical datasets.

**Methods:** Each dataset was split into five folds for cross-validation. Afterwards, a hybrid dataset was created to test for transfer learning. We tested three classic machine learning models and a custom deep learning model that were trained using electronic healthcare record (EHR) data from each patient. We evaluated each model for predictive performance at 60, 120, 180, and 365 days ahead of amputation in the *All of Us* data set. We then evaluated model generalizability by evaluating model performance on an institutional data set *STARR*.

**Results:** Our deep learning model had average AUCs of 0.90 (60 days prior to amputation) and 0.89 (120, 180 and 365 days prior to amputation) in the *All of Us* data set. Models were then externally validated on the *STARR* dataset, achieving average AUC scores of 0.73 (60 days), 0.71 (120 and 365 days), 0.70 (180 days).

**Conclusions:** Our prognostic deep learning model performs well in predicting risk of amputation up to a year prior. This model also demonstrates promising generalizability by showing strong performance during external validation on a local, institutional dataset. Our deep learning model also adapted the best to the hybrid dataset.

## Introduction

Peripheral artery disease (PAD) is associated with 51%-93% of amputations depending on geographic region.^1^ Major amputation is associated with high mortality—12.4% within 30 days of amputation.^2^ Contemporary medical and surgical techniques for averting limb loss in individuals with PAD demonstrate remarkable efficacy. Employing antiplatelet agents, statin therapy, supervised exercise, guidance for quitting smoking, and managing underlying conditions like hypertension and diabetes substantially diminishes the likelihood of major adverse cardiovascular and limb-related incidents.^3^ Implementing revascularization methods, including surgical bypass to redirect blood circulation to the lower limbs, alongside innovative minimally invasive endovascular procedures, has notably decreased amputation rates in PAD patients. Barnes, et al. recently reviewed the epidemiology and risk of amputation in patients with diabetes mellites and PAD, finding that patients with both conditions face significantly higher risks of amputation than patients with either disease by itself.^4^

While evidence-based strategies to combat amputation are known, there are no widely adopted risk models that can identify patients at highest risk of amputation^5^ Cox, et al. analyzed a dataset of 14,444 patients that underwent lower extremity endovascular procedures, and were able to predict 30-day amputation using a random forest model with an AUC-ROC score of 0.81.^6^ Kreutzburg, et al. used a Cox proportional hazards model to predict five year survival (AUC of 0.70) and major amputation above ankle level (AUC of 0.69) within at least one month after discharge.^7^ More generally, Ross, et al. analyzed EHR data with lab results to build random forest and linear regression models to predict major adverse cardiac and cerebrovascular events after PAD diagnosis, with their best model achieving an AUC of 0.81.^8^ Liu, et al. analyze patient histories and labs to predict amputation-free survival on a structured dataset after revascularization in patients with PAD attaining an AUC of 0.82, 0.80, and 0.80 for one, three, and five year amputation-free survival using a random survival forest.^9^. Thus, there remains a need to develop a system that can provide early alerts so that high risk patients can be more aggressively managed medically and/or surgically.

In this study, we build and compare multiple machine learning models to predict amputation in patients with PAD. We look at a wide variety of traditional machine learning algorithms and a custom deep learning algorithm that fuses sequential histories and structured demographic data. Additionally, we explore model generalizability. In the past years, there has been a lot of debate and investigation about the generalizability of machine learning across clinical contexts.^10,11^ Specifically, the question of generalizability is important, as well as whether models can work across clinical settings with different distributions. Thus, we employ two different datasets of electronic healthcare records to assess the generalizability of our models.

## Methods

To train and validate our models, we used deidentified EHR spanning all observations, conditions, procedures performed, and prescribed medications and demographic information for our two pre-defined PAD cohorts. We constructed three different configurations of internal and external testing: First, we performed 5-fold cross validation training and tested on the *All of Us* dataset, with each fold externally tested on the *STARR* dataset. Secondly, we also trained and tested on just the *STARR* dataset to have a baseline for comparison. Finally, we trained and tested on a hybrid dataset consisting of the *All of Us* dataset supplemented with 50% of the smaller *STARR* dataset, with each fold tested on the remaining 50% of the *STARR* dataset in order to test transfer learning. All models were coded in Python (v 3.10.12). Scikit Learn (v1.3.0) was used for preprocessing, standardization, and the traditional machine learning models.^12^ Keras (v2.11.0) was used for deep learning architecture.^13^ To collect the data for All of Us we used the R package allofus.^14^ We constructed models with different follow-up times for the event: 60 days (2 months), 120 days (4 months), 180 days (6 months), and 365 days (1 year). Since patients with gangrene conditions are already closely monitored for amputation risk, we eliminated these patients from the cohort. To guarantee sufficient quality of data, we also imposed a minimum length of observation of at least two weeks and 20 recorded observations. These limitations significantly lower the number of patients with amputations included in our models depending on the follow-up time.

### Data Sources

We employed two datasets from different sources. The first dataset derived from the Stanford Medicine Research Data Repository (*STARR*). Data includes de-identified EHR clinical practice data at Stanford Medicine from 1998 to 2022. It consists of 2,437,028 clinical features for treatments, diagnoses, and other medical concepts, with 6,539 unique patients diagnosed with PAD with 312 having undergone a non-traumatic amputation. Use of the STARR dataset for this study was approved by the Stanford IRB. The second dataset is derived from the *All of Us* program, which started enrollment in May of 2018 and contains a variety of genetic, survey, lab, and EHR data for a large and diverse cohort of volunteers from the United states.^15^ Our *All of Us* (Registered Tier v7) dataset consisted of 29,663,453 patient clinical features from 10,153 unique patients, of which 710 underwent a non-traumatic amputation. The Registered Tier and Controlled Tier data available on the Research Hub contains data from participants who have consented to be involved in the All of Us Research Program, including data from electronic health records (EHRs), surveys, and physical measurements. All data available to researchers has had direct identifiers removed and has been further modified to minimize re-identification risks.

To select PAD patients, we used the PAD conditions named with at least one SNOMED/ICD9 code associated with peripheral artery disease, and we selected amputations through a list of SNOMED codes of non-traumatic amputations. Note that for amputation cases, patient histories are taken to the point of amputation.

### Machine Learning and Deep Learning Models

For our traditional machine learning models, we took the electronic health care record data and encoded each concept by the number of times it occurs as a code in a patient’s records. We evaluated and compared three different models: random forest (RF), logistic regression (LR), and XGBoost (XGB).

For our deep learning model (Figure 1), we utilized the model described by Ghanzouri, et al for classifying PAD patients. ^16^ The model consists of two separate blocks. For the sequential EHR data we used a long short-term memory recurrent neural net (LSTM) model, and for the static demographic features, we used a parallel fully connected neural network. We call this combined model LSTM+. The two substructures of our deep learning model are then combined using late fusion into a binary classifier for amputation or non-amputation. Standard rectified linear unit (ReLU) functions were used throughout the model except for a sigmoid function as the final layer of the binary classifier. Model weights were updated through training using the Adaptive Moment Estimation (Adam) optimizer. From manual parameter searching, we chose the best hyperparameters for the 60-day model trained on *All of Us* and kept those hyperparameters throughout the rest of the models. We tested models across the following metrics: Area Under the Receiver Operator Curve (AUC), Specificity, Sensitivity (SEN), Positive Predictive Value (PPV), Negative Predictive Value (NPV) at optimal thresholds (see Supplemental Tables I-III for full metrics). Notably, the relative rarity of amputations means that the positive predictive value is the most affected metric due to the class imbalance.

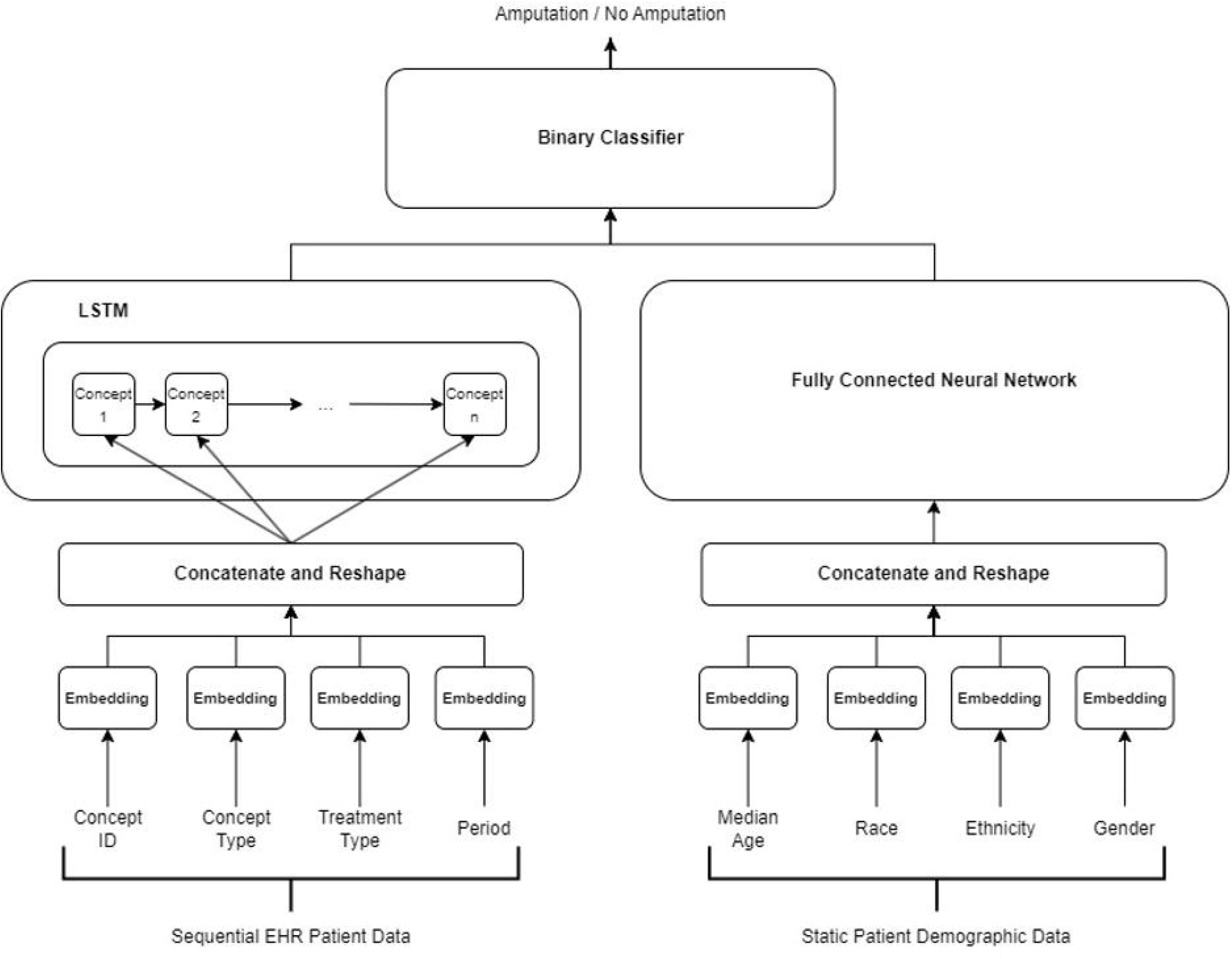

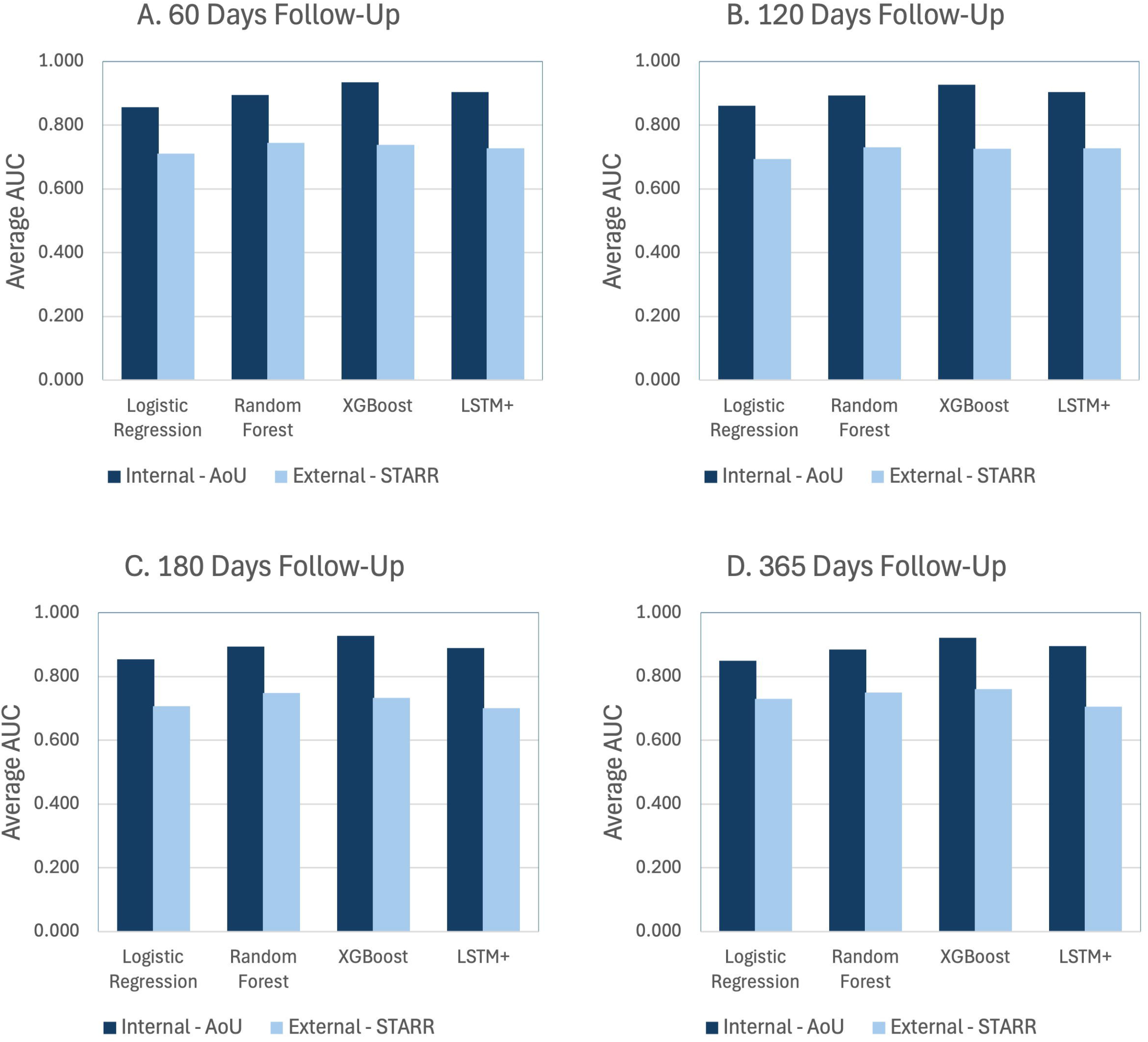

## Results

See Table I for a descriptive summary of the cohort. Mean age (years) is calculated from the age at first diagnosis of a PAD condition. As described in Table I, there are significant differences in demographic and clinical comorbidities across amputation cases and controls. For example, there is a difference between the mean age in cases versus controls across both *All of Us* and *STARR*, with a mean age at PAD diagnosis of 54.5 years in amputation cases in *STARR* versus 66.3 years in the non-amputation controls. In *All of Us*, the mean age at PAD diagnosis for patients with amputations is 63.3 years versus 70.3 years for those in the non-amputation control. Notably, there is also a significant difference in the length of history between cases and controls with an average observation length of EHR data of 763 days for patients with amputations in *STARR* compared to 2370.6 days for control patients. In *All of Us* the average observation length for amputation cases is 1755.7 days compared to 6428.1 days for patients without amputations. This shows a considerable class imbalance in the length of patient histories, which is reflected in some of the rates of health disorders observed, e.g. heart failure that manifests after amputation is not included in our counts. Furthermore, all comorbidities were found at higher rates in *STARR* dataset compared to the *All of Us* dataset.

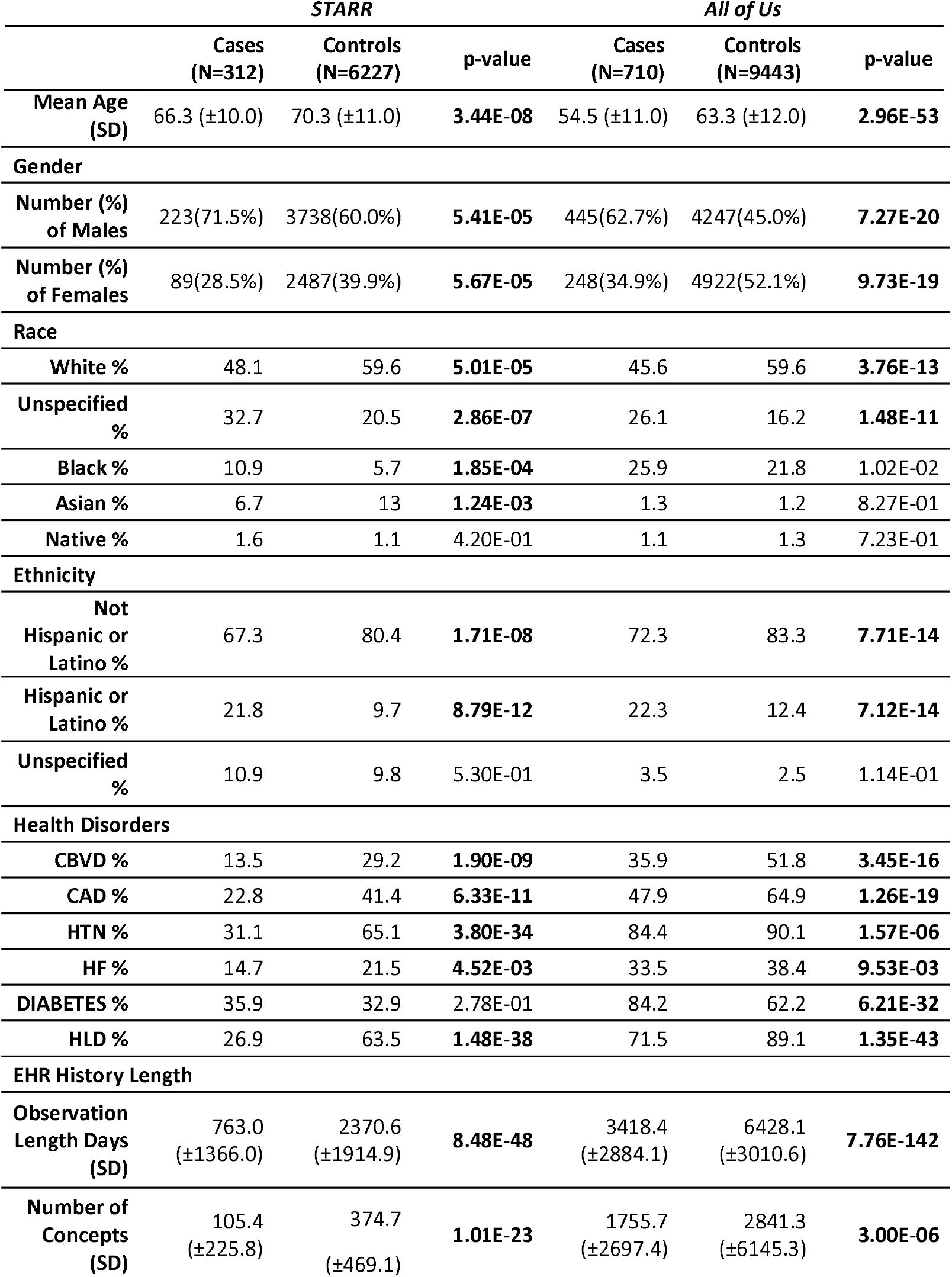

| *STARR* |  |  | *All of Us* |  |  |
| --- | --- | --- | --- | --- | --- |
| Cases<br>(N=312) | Controls<br>(N=6227) | p-value | Cases<br>(N=710) | Controls<br>(N=9443) | p-value |

|  |  |  |  |  |  |  |
| --- | --- | --- | --- | --- | --- | --- |
| **Mean Age (SD)** | 66.3 (±10.0) | 70.3 (±11.0) | **3.44E-08** | 54.5 (±11.0) | 63.3 (±12.0) | **2.96E-53** |
| **Gender** |  |  |  |  |  |  |
| **Number (%) of Males** | 223(71.5%) | 3738(60.0%) | **5.41E-05** | 445(62.7%) | 4247(45.0%) | **7.27E-20** |
| **Number (%) of Females** | 89(28.5%) | 2487(39.9%) | **5.67E-05** | 248(34.9%) | 4922(52.1%) | **9.73E-19** |
| **Race** |  |  |  |  |  |  |
| **White %** | 48.1 | 59.6 | **5.01E-05** | 45.6 | 59.6 | **3.76E-13** |
| **Unspecified %** | 32.7 | 20.5 | **2.86E-07** | 26.1 | 16.2 | **1.48E-11** |
| **Black %** | 10.9 | 5.7 | **1.85E-04** | 25.9 | 21.8 | 1.02E-02 |
| **Asian %** | 6.7 | 13 | **1.24E-03** | 1.3 | 1.2 | 8.27E-01 |
| **Native %** | 1.6 | 1.1 | 4.20E-01 | 1.1 | 1.3 | 7.23E-01 |
| **Ethnicity** |  |  |  |  |  |  |
| **Not Hispanic or Latino %** | 67.3 | 80.4 | **1.71E-08** | 72.3 | 83.3 | **7.71E-14** |
| **Hispanic or Latino %** | 21.8 | 9.7 | **8.79E-12** | 22.3 | 12.4 | **7.12E-14** |
| **Unspecified %** | 10.9 | 9.8 | 5.30E-01 | 3.5 | 2.5 | 1.14E-01 |
| **Health Disorders** |  |  |  |  |  |  |
| **CBVD %** | 13.5 | 29.2 | **1.90E-09** | 35.9 | 51.8 | **3.45E-16** |
| **CAD %** | 22.8 | 41.4 | **6.33E-11** | 47.9 | 64.9 | **1.26E-19** |
| **HTN %** | 31.1 | 65.1 | **3.80E-34** | 84.4 | 90.1 | **1.57E-06** |
| **HF %** | 14.7 | 21.5 | **4.52E-03** | 33.5 | 38.4 | **9.53E-03** |
| **DIABETES %** | 35.9 | 32.9 | 2.78E-01 | 84.2 | 62.2 | **6.21E-32** |
| **HLD %** | 26.9 | 63.5 | **1.48E-38** | 71.5 | 89.1 | **1.35E-43** |
| **EHR History Length** |  |  |  |  |  |  |
| **Observation Length Days (SD)** | 763.0 (±1366.0) | 2370.6 (±1914.9) | **8.48E-48** | 3418.4 (±2884.1) | 6428.1 (±3010.6) | **7.76E-142** |
| **Number of Concepts (SD)** | 105.4 (±225.8) | 374.7 (±469.1) | **1.01E-23** | 1755.7 (±2697.4) | 2841.3 (±6145.3) | **3.00E-06** |

Figure 2 shows our results for the first configuration, when the models are trained on the *All of Us* dataset then tested both internally and externally on the *STARR* dataset.

For 60 follow-up predictions, XGBoost has the best average AUC of 0.935 on internal *All of Us* testing; however, notably our LSTM+ has the highest PPV 0.313. For the external testing at 60 days follow-up, Random Forest proves robust with an AUC of 0.744. As we increase the number of days, we show several different follow-up intervals with different time periods.

In Figure 3, we provide the performance of our models on just *STARR* dataset. Since this is a smaller dataset with a much smaller number of cases, we can expect weaker performance overall, particularly with the ability of the models to accurately identify cases. Our LSTM+ architecture proves surprisingly robust for the *STARR* dataset, with an AUC of 0.871 for predicting 60-day follow-up. Even at the longer time frames, LSTM+ remains the strongest model overall. With the shorter time series and observation lengths of the *STARR* dataset, it seems the ability of our LSTM+ to incorporate the sequential structures of the EHR data improves its comparative performance.

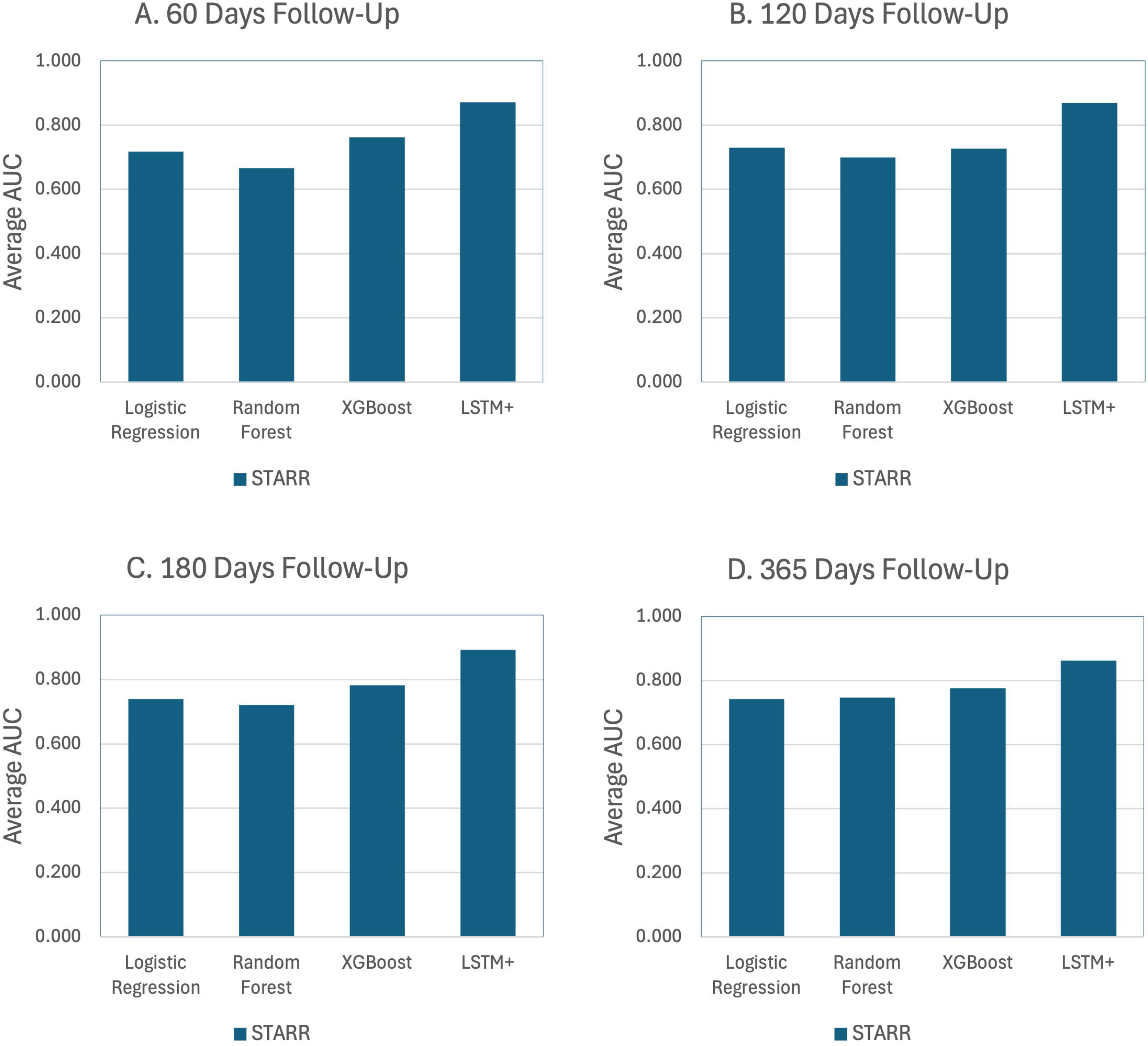

In Figure 4, we present our hybrid training strategies, where half of the *STARR* dataset was combined with the *All of Us* dataset for five-fold internal cross-validation. The remaining half of the *STARR* dataset is withheld for external testing.

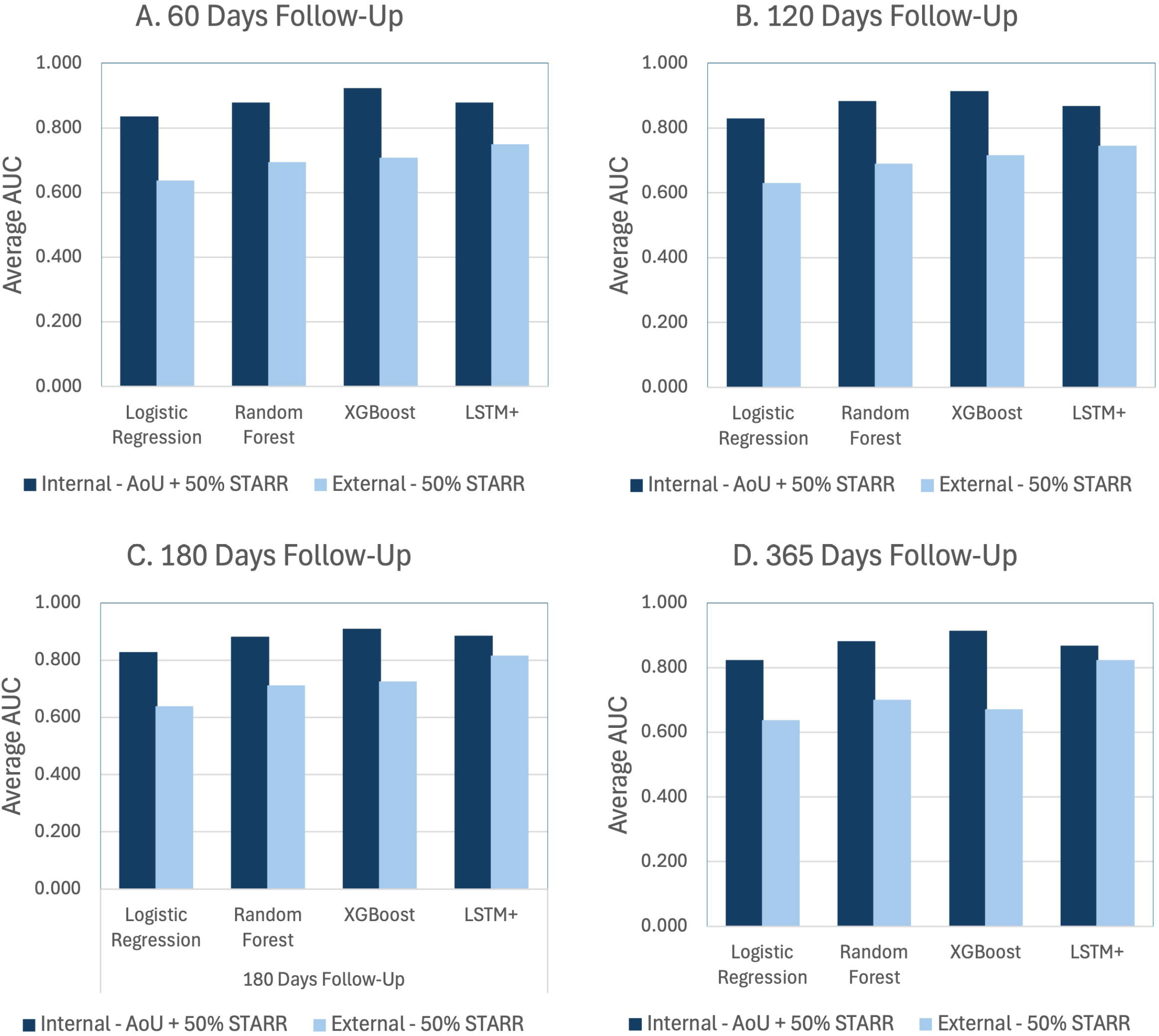

As in the *All of Us* trained case, XGBoost has the most robust internal testing metrics. However, LSTM+ proves to have stronger external testing performance on the remaining half of the *STARR* dataset, with an external average AUC of 0.75, 0.75, 0.82, and 0.82 at 60, 120, 180, and 365 days follow-up respectively.

In Supplemental Tables IV-V, we include two tables of the top twenty-five features by importance from XGBoost for both the *All of Us* and *STARR* datasets. Notably, ulcer of the foot is the top feature across all time frames and both datasets, which implies that although foot ulcers are an important predictor of years before amputation occurs. Also, tobacco use as a feature appears mostly in the 365-days follow up models, but not in the shorter follow-up periods. Various pain conditions of the lower body (hip, knee, and back) seemed to be strong predictors in the *All of Us* dataset, but not in the *STARR* dataset. Unfortunately, there are no effective methods to measure the importance of a feature in our LSTM model architecture.

## Discussion

We demonstrate that as expected, XGBoost was generally better able to manage the class imbalance in our datasets than logistic regression or random forest.^17^ While XGBoost had better performance on the *All of Us* trained models on internal testing, our deep learning LSTM+ had much better performance on the smaller *STARR* dataset. Furthermore, the hybrid datasets show that the LSTM+ also has considerably better performance on external testing on the *STARR* dataset. In general, traditional machine learning model performance decreased over longer follow-up time periods. The performance of the LSTM+ improves as the time frame of prediction increases is interesting, but most likely indicates that as we increase the number of days we predict ahead, we naturally eliminate patients with shorter histories and tend to select patients with longer and more robust histories in the smaller *STARR* dataset. This indicates that our deep learning architecture can better adapt to smaller datasets with different distributions.

There have been only a few recent studies using artificial intelligence to detect and analyze patients with PAD. In their literature review, Flores et al. identify only three papers that use EHR data to build models that diagnose PAD before 2021.^18^ Ross et al. compared several classic machine learning models to classify PAD in a dataset of 1,755 patients who presented for elective coronary angiography and found that a penalized regression model performed the best with an AUC-ROC score of 0.87.^19^ Zhang, et al. compared various machine learning approaches to predict in-hospitality mortality for patients identified with PAD.^20^ Ghanzouri, et al. developed an automated tool to identify undiagnosed PAD, and achieved an AUC score of 0.96 with a deep learning model using a dataset of 3,168 patients.^16^

Very few papers use EHR data to predict adverse events in PAD patients. Davis, et al. used the Super Learner ensemble-model to predict surgical site infection in patients with PAD after open lower extremity bypass. Our study is not limited to patients to tracking patients after a procedure while having the ability to assess risk at four different time periods. While it is difficult to compare directly, the performance of our models compares favorably to the other works in the field of predicting amputations. Kreutzburg, et al. had an AUC of 0.69 for one-year amputation after a month of discharge.^7^ Liu, et al. specifically looked at amputation free survival after successful revascularization in patients with PAD, a much narrower cohort of 1,260 patients, had a one-year amputation free survival AUC of 0.821.^9^ Our LSTM+ model with predicting more than 60 days follow-up, had an AUC of 0.903 on the *All of Us* dataset and an AUC of 0.871 on the *STARR* dataset. To the best of our knowledge, our work is the only study on Machine Learning for PAD that tests our models on more than one dataset and its ability to adapt to new domains; furthermore, our study is the first to build models for four different time periods of possible intervention.

## Limitations

Our study has a few limitations. Due to the relative rarity of amputations in our datasets, our positive predictive value is relatively limited, despite the otherwise strong metrics. Furthermore, we eliminate patients with histories of short lengths of less than two weeks, it may be difficult to implement in clinical contexts where full patient histories are not available. Our deep learning model cannot score feature importance, so it lacks the ability to explain why a patient may be at risk.

## Conclusions

We used two different large clinical populations ( and *STARR*) to develop a robust deep learning model across four different time frames to predict amputation events in PAD patients. We achieved strong AUC performance across both datasets and showed the ability of our models to adapt to different domains. Our models can be implemented to help inform clinicians of potential risk of amputation in patients with PAD.

## Supporting information

Supplemental Table III

## Data Availability

The cohorts constructed from the datasets are available upon reasonable request to the authors for researchers that have the appropriate access to each dataset.

https://med.stanford.edu/starr-tools.html

https://allofus.nih.gov/

## Acknowledgments

We thank the Institute for Experiential AI at Northeastern University for their support and the Harold Alfond foundation for making the Roux Institute possible. We gratefully acknowledge All of Us participants for their contributions, without whom this research would not have been possible. We also thank the National Institutes of Health’s All of Us Research Program for making available the participant data examined in this study. This research used data or services provided by STARR, “STAnford medicine Research data Repository,” a clinical data warehouse containing live Epic data from Stanford Health Care, the Stanford Children’s Hospital, the University Healthcare Alliance and Packard Children’s Health Alliance clinics and other auxiliary data from Hospital applications such as radiology PACS. STARR platform is developed and operated by Stanford Medicine Research Technology team and is made possible by Stanford School of Medicine Research Office. We thank Michael Wilczek, Ilies Ghanzouri for helping in collecting and constructing the datasets, and Naim Shehadeh, Nawar Shara for helping with the conception and design of the research.

